# Increased prolonged sitting in rheumatoid arthritis patients during the COVID-19 pandemic: a within-subjects, accelerometer-based study

**DOI:** 10.1101/2020.09.09.20191395

**Authors:** Ana J. Pinto, Diego Rezende, Sofia M. Sieczkowska, Kamila Meireles, Karina Bonfiglioli, Ana C. M. Ribeiro, Eloisa Bonfá, Neville Owen, David W. Dunstan, Hamilton Roschel, Bruno Gualano

**Affiliations:** Applied Physiology and Nutrition Research Group; Laboratory of Assessment and Conditioning in Rheumatology; Hospital das Clínicas HCFMUSP, Faculdade de Medicina FMUSP, Universidade de Sao Paulo – Av. Dr. Arnaldo, 455, ZIP code: 01246-903, Sao Paulo-SP, Brazil; Rheumatology Division, Faculdade de Medicina FMUSP, Universidade de Sao Paulo – Av. Dr. Arnaldo, 455, ZIP code: 05403-900, Sao Paulo-SP, Brazil; Baker Heart and Diabetes Institute, Melbourne VIC, Australia – 99 Commercial Road, Melbourne, Victoria 3004, Australia; Centre for Urban Transitions, Swinburne University of Technology – John St, Melbourne, Victoria 3122, Australia; Mary McKillop Institute for Health Research, Australian Catholic University – 215 Spring St, Melbourne, Victoria 3000, Australia; Food Research Center, University of Sao Paulo – R. do Lago, 250, ZIP code: 05508-080, Sao Paulo-SP, Brazil

**Author notes:** Corresponding author: Prof. Dr. Bruno Gualano; Applied Physiology & Nutrition Research Group. Rheumatology Division, Faculdade de Medicina FMUSP, Universidade de São Paulo – Av. Dr. Arnaldo, 455, 3º andar, ZIP code: 01246–903, Sao Paulo – SP, Brazil.

**Keywords:** physical activity, sedentary behavior, inflammatory arthritis

## Abstract

**Background:** Social distancing measures designed to contain the COVID-19 pandemic can explicitly and implicitly restrict physical activity, a particular concern for high-risk patient groups. Using a within-subjects design with objective measurement (via validated accelerometers), we assessed rheumatoid arthritis patients’ physical activity and sedentary behavior levels prior to and during the social distancing measures implemented in Sao Paulo, Brazil.

**Methods:** Post-menopausal women diagnosed with rheumatoid arthritis were assessed before (from March 2018 to March 2020) and during (from 24^th^ May to 7^th^ July 2020) social distancing measures to contain COVID-19 pandemic, using a within-subjects, repeated-measure design. Physical activity and sedentary behavior were assessed using postural-based accelerometry (ActivPAL micro™).

**Findings:** Mean age was 60.9 years (95%CI: 58.0, 63.7) and BMI was 29.5 Kg/m^2^ (95%CI: 27.2, 31.9). Disease activity ranged from remission to moderate activity. Most of the patients were using disease-modifying anti-rheumatic drugs and prednisone. Hypertension and dyslipidemia were the most frequent comorbidities During social distancing, there were reductions in total stepping time (15.7% [−0.3 h/day, 95%CI: −0.4, −0.1; p = 0.004]), in light-intensity activity (13.0% [−0.2 h/day, 95%CI: −0.4, −0.04; p = 0.016]) and in moderate-to-vigorous physical activity (38.8% [−4.5 min/day, 95%CI: −8.1, −0.9; p = 0.015]), but no changes in total standing time or total sedentary time. However, time spent in prolonged bouts of sitting ≥ 30 min increased by 34% 1.0 h/day, 95%CI: 0.3, 1.7; p=0.006) and sitting bouts ≥60 min increased by 85% (1.0 h/day, 95%CI: 0.5, 1.6) Sit-stand transitions were reduced by 10% (−5.1/day, 95%CI: −10.3, 0.0; p = 0.051)

**Conclusion:** Imposed social distancing measures to contain the COVID-19 outbreak were associated with decreased physical activity and increased prolonged sitting among rheumatoid arthritis patients. Since this has the potential to increase the burden of cardiovascular disease in such high-risk patients, attention to maintaining physical activity is an urgent consideration during the pandemic.

## Introduction

A preliminary, multinational survey reporting step counts provided by smartphones showed that social distancing measures to contain the spread of SARS-CoV-2 have induced physical inactivity (i.e., not meeting the physical activity guidelines) (1), which is strongly associated with morbidity and all-cause mortality (2). Sedentary behavior (i.e., time spent in a sitting or reclining posture) is also related to poor health outcomes that are in addition to inactivity (3), but the impact of social distancing on sedentary behavior during the pandemic is currently unknown.

Physical inactivity and sedentary behavior are modifiable risk factors considered to be potential targets to prevent morbimortality in autoimmune rheumatic diseases (4, 5). Among rheumatoid arthritis (RA) patients, sedentary behavior is associated with higher disease scores, increased pain, fatigue (6) and number of comorbidities, reduced aerobic capacity (7) and physical function (6), and poor self-efficacy (8). Furthermore, physically inactive RA patients exhibit higher cardiovascular risk factors (e.g., higher systolic blood pressure and HOMA index, abnormal lipid profile) when compared to their physically active counterparts.

RA patients have been shown to be more susceptible to COVID-19 infection(9) and, therefore, may be subjected to more restrictive measures of social distancing, potentially with significant impacts on their activity options, and, hence, on their burden of cardiovascular disease risk, the main cause of mortality in this population (10).

In this prospective study using a within-subjects design, we assessed RA patients’ physical activity and sedentary behavior levels using accelerometers, prior to and during the imposed measures of social distancing to combat COVID-19 in Sao Paulo, Brazil.

## Materials and Methods

### Sample and experimental design

Sixty-four post-menopausal women diagnosed with RA (11) were recruited from the Outpatient Rheumatoid Arthritis Clinic of the Clinical Hospital (School of Medicine, University of Sao Paulo) between March 2018 and March 2020 to participate in a trial (clinicaltrials.gov: NCT03186924). At baseline, patients were not engaged in exercise training programs within the last 12 months and had stable drug therapy in the last three months. This trial was approved by the local ethical committee (Commission for Analysis of Research Projects). Patients signed an informed consent form before participation. All patients had been through a clinical and physical activity assessment before the official set of social distancing measures to contain the COVID-19 outbreak, adopted on the 24^th^ of March. This facilitated the unique opportunity to track physical activity levels during the pandemic in a within-subjects, repeated measure design. We then obtained a new approval from the ethics committee for collecting data during the social distancing. Thirty-five out of 64 patients accepted to participate. Three members of our staff (DR, SMS, KM) delivered the accelerometers (ActivPAL micro™, PAL Technology, UK) to the patients at home from the 24^th^ May to 7^th^ July. The time elapsed for data collection between baseline and during social distancing was 12.5 months (9.9, 15.2). Patients were asked if they had adhered to the social distancing measures. All but two responded affirmatively. Data were assessed with and without the two non-compliers, and results remained the same. Thus, we reported the full data.

### Accelerometry

Patients were instructed to wear the accelerometer on the medial front of the right thigh during 7 days for 24 h. Data were exported and analyzed using ActivPAL3™ software, version 8.10.9.46 (PAL Technology, UK). Data was checked by an experienced researcher and also crosschecked with a sleep diary. Time spent in moderate-to-vigorous physical activity was calculated as time spent above a step cadence of ≥ 100 steps/min (12).

### Statistics

Dependent variables were tested using repeated measures mixed models, with time as fixed factor and participants as random factor, using SAS 9.3 (SAS Institute Inc., Cary, USA). Data are presented as mean (95%CI). The significance level was P≤0.05.

## Results

Mean age was 60×9 years (95%CI: 58.0, 63.7) and BMI was 29.5 Kg/m^2^ (95%CI: 27.2, 31.9). Disease activity ranged from remission to moderate activity. Most of the patients were using disease-modifying anti-rheumatic drugs and prednisone. Hypertension and dyslipidemia were the most frequent comorbidities (Table 1).

**Table 1.** Patients’ characteristics prior to the COVID-19 pandemic.

| Variable | n=35 |
| --- | --- |
| Age, years | 60.9 (58.0, 63.7) |
| BMI, kg/m<sup>2</sup> | 29.5 (27.2, 31.9) |
| Disease Activity Score (DAS28 PCR) | 3.4 (2.9, 3.8) |
| Clinical Disease Activity Index (CDAI) | 10.1 (6.7, 13.4) |
| Health Assessment Questionnaire | 1.2 (0.9, 1.4) |
| C-reactive protein, mg/L | 10.8 (5.5, 16.2) |
| Erythrocyte sedimentation rate, mm/H | 28.4 (15.7, 41.1) |
| Disease duration, years | 18.5 (14.7, 22.3) |
| Comorbidities, n (%) |  |
| Hypertension | 18 (51.4%) |
| Dyslipidemias | 17 (48.6%) |
| Type 2 diabetes | 12 (34.3%) |
| Fibromyalgia | 11 (31.4%) |
| Other rheumatic diseases | 18 (51.4%) |
| Depression | 7 (20.0%) |
| Medication, n (%) |  |
| DMARDs | 30 (85.7%) |
| Leflunomide | 19 (54.3%) |
| Methotrexate | 16 (45.7%) |
| Prednisone | 26 (74.3%) |
| Biological agents | 12 (34.3%) |
| Anti-inflammatory drugs | 11 (31.4%) |
| Pain killers | 20 (57.1%) |
| Antihypertensive drugs | 18 (51.4%) |
| Antihyperlipidemic drugs | 17 (48.6%) |
| Antidiabetic drugs | 12 (34.3%) |
| Anti-depressants | 14 (40.0%) |

Data presented as mean (95%CI) or n (%). Other rheumatic diseases include
osteoarthritis, osteoporosis or Sjogren's syndrome. Abbreviations: DMARDs,
disease-modifying antirheumatic drugs; TNF, tumor necrosis factor.

During social distancing, there were reductions in total stepping time (15.7% [-0.3 h/day, 95%CI: −0.4, −0.1; p = 0.004]), in light-intensity activity (13.0% [-0.2 h/day, 95%CI: −0.4, −0.04; p = 0.016]) and in moderate-to-vigorous physical activity (38.8% [-4.5 min/day, 95%CI: −8.1, −0.9; p = 0.015]), but no changes in total standing time (−0.1 h/day, 95%CI: −0.7, 0.5; p = 0.767) or total sedentary time (0.3 h/day, 95%CI: −0.4, 1.0; p = 0.335).

However, time spent in prolonged bouts of sitting ≥30 min increased by 34% (p = 0.006; Figure 1, panel A) and sitting bouts ≥60 min increased by 85% (p = 0.001; Figure 1, panel B). Sit-stand transitions were reduced by 10% (−5.1/day, 95%CI: −10.3, 0; p = 0.051).

**Figure 1.**
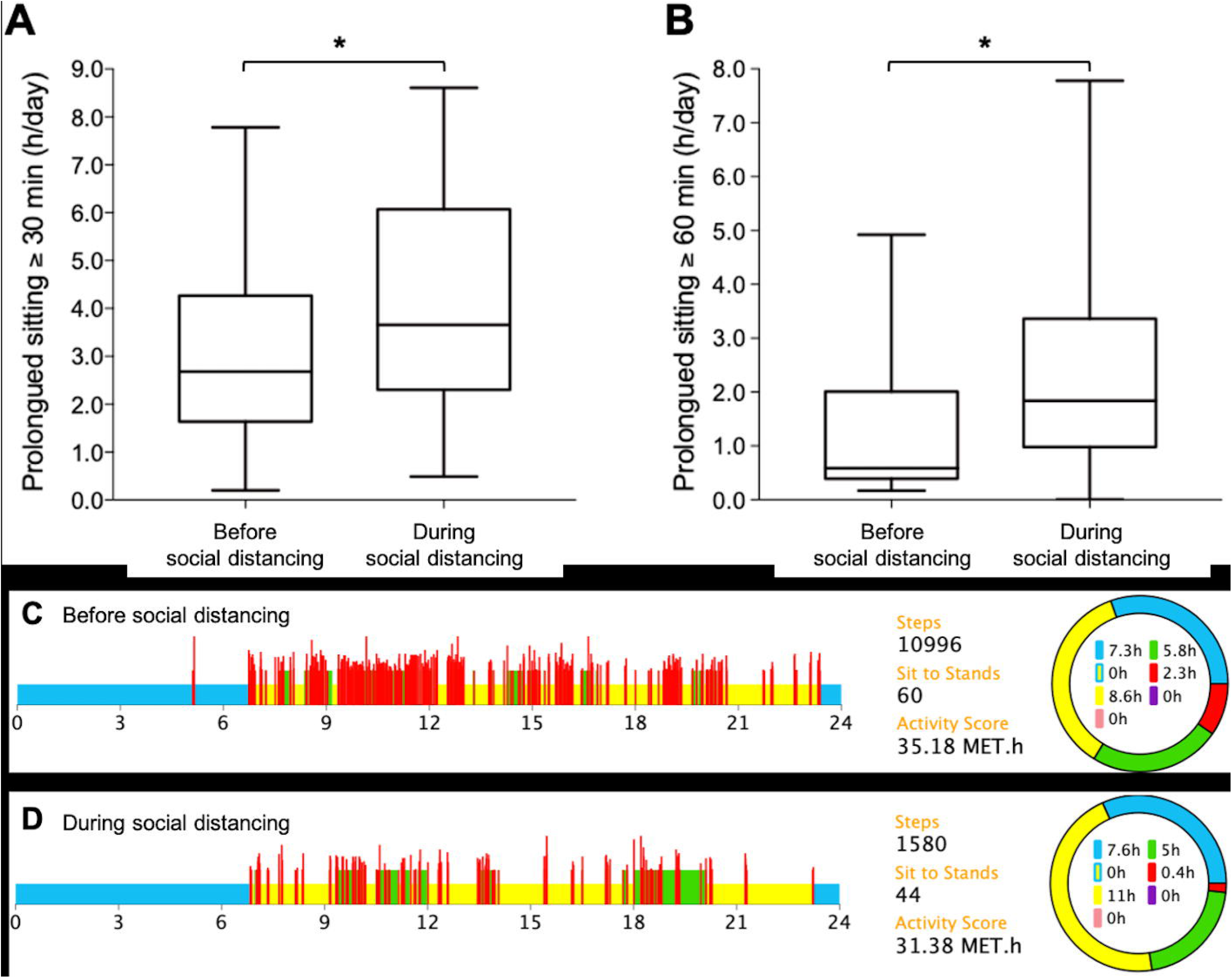
Prolonged sitting and physical activity in rheumatoid arthritis. Panels A and B depict total time spent in prolonged sitting ≥30 min and ≥60 min. In the bottom, illustrative data showing the fall in activity and the rise in prolonged sitting in a RA patient assessed before (Panel C) and during the social distancing (Panel D). Legend: blue, sleeping time; yellow, sedentary time; green, standing time; red, stepping time (irrespective of intensity); *p<0.05.

Panels C and D illustrate the accelerometer data from a patient who experienced decreased activity and increased prolonged sitting after social distancing.

## Discussion

To our knowledge, this is the first study to track physical activity and sedentary behavior patterns before and during the COVID-19 pandemic using validated accelerometers and a within-subjects design. Our main findings suggest that social distancing (including stay-at-home order) can lead to *i)* increased physical inactivity, and, additionally, *ii)* increased prolonged sitting among RA patients. Inactivity along with too much sitting emerge as a risk factor that could be detrimental to overall health in such a high-risk group of patients during the COVID-19 pandemic.

Observational and experimental evidence demonstrates that inactivity can predispose to pathological states and poor outcomes (13). Sedentary behavior can add to the adverse impacts of physical inactivity in impairing cardiovascular health (14). Autoimmune rheumatic patients commonly spent most of their daily hours engaged in sedentary behavior and did not achieve minimum levels of moderate-to-vigorous physical activity (5).

Namely in RA, the estimates of physical inactivity and sedentary behavior are comparable to those of other chronic diseases (e.g., type 2 diabetes and cardiovascular diseases) and associate with poor health-related outcomes (5). As those confined at home are less prone to perform physical activity, it has been speculated that inactivity and sedentary behavior could peak during the COVID-19 pandemic (4). This is of particular concern for those who are usually hypoactive and show higher risk of cardiovascular diseases, this being the case of RA patients (see the patients’ comorbidities in Table 1) (5).

Interestingly, even in the absence of changes in total sedentary time, prolonged sitting time rose considerably. Uninterrupted sedentary bouts are associated with all-cause mortality (15), whereas well-controlled studies show that very-light active interruptions in prolonged sedentary time (e.g., 2 minutes of walking for every 30 minutes of sitting) can elicit immediate improvements in postprandial metabolism, blood pressure, and vascular function (16–18).

This raises the need for widespread recommendation of breaking-up prolonged sitting whenever possible (e.g., 2-min breaks of walking every 30 min of sitting) to avoid poor outcomes during the social distancing rules, which tend to be more restrictive for high-risk groups for COVID-19, such as those with autoimmune rheumatic diseases (9).

The main strengths of this study are its within-subjects design and the use of posture-based accelerometers, which enables an objective and a comprehensive assessment of sedentary behavior patterns. The limitations include the relatively low sample size and the inability to stablish a cause-andeffect relationship between changes in behavior with social distancing measures, although elements of temporality and plausibility do support our assumptions.

In conclusion, imposed social distancing measures to contain the COVID-19 outbreak were associated with decreased physical activity and increased prolonged sitting time among RA patients. Since this has the potential to increase the burden of cardiovascular diseases in such high-risk patients, attention to maintaining their activity levels is an urgent consideration during the pandemic.

## Data Availability

Data will be available upon reasonable request.

## Acknowledgements

AJP, SMS and BG were supported by Fundação de Amparo à Pesquisa do Estado de São Paulo (FAPESP, 2015/26937-4; 2019/15231-4; 2017/13552-2). DR and KM were supported by Coordenação de Aperfeiçoamento de Pessoal de Nível Superior (CAPES) − Finance Code 001.

## Conflict of interests

The authors declare no conflict of interests.

## Author contributions

Conception and design: AJP, BG; participants’ recruitment and data collection: AJP, DR, SMS, KM; data analysis and interpretation: AJP, KB, ACMR, EB, NO, DWD, HR, BG; manuscript writing: AJP, BG; manuscript revision DR, SMS, KM, KB, ACMR EB, NO, DWD, HR. All authors have approved the final version.

## Notes

### Competing Interest Statement

The authors have declared no competing interest.

### Funding Statement

AJP, SMS and BG were supported by Fundacao de Amparo a Pesquisa do Estado de Sao Paulo (FAPESP, 2015/26937-4; 2019/15231-4; 2017/13552-2). DR and KM were supported by Coordenacao de Aperfeicoamento de Pessoal de Nivel Superior (CAPES), Finance Code 001.

### Author Declarations

This trial was approved by the local ethical committee (Commission for Analysis of Research Projects).

